# Investigating associations between hearing, cognition, and social isolation using the Hertfordshire Ageing Study

**DOI:** 10.1101/2025.07.02.25330728

**Authors:** Nisha Dhanda, Amanda Hall, James Martin, Helen Pryce

**Affiliations:** Department of Applied Health Sciences, College of Medicine and Health, University of Birmingham, Edgbaston UK; Department of Audiology, College of Health and Life Sciences, Aston University, Birmingham UK

**Keywords:** epidemiology, hearing, cognition, social isolation, older adults, ageing

## Abstract

**Objectives:**

1. To investigate whether hearing threshold predicts cognitive score 10 years later using the Hertfordshire Ageing Study data.
2. To investigate whether hearing threshold predicts social isolation score 10 years later using the Hertfordshire Ageing Study data.

**Methods:** The Hertfordshire Ageing Study (HAS) is a longitudinal cohort study that measured hearing via pure tone audiometry at two time points, and social isolation and cognition variables at the second time point. Linear regression was implemented for both objectives using an unadjusted model, a model controlling for age and gender, and a model controlling for all confounders (sociodemographic, lifestyle and clinical characteristics) relating to the exposure and outcome variables.

**Results:** 231 participants were included in the final analysis. There was no statistically or clinically significant evidence found to indicate hearing threshold at timepoint 1 predicting cognitive score at timepoint 2, and hearing threshold at timepoint 1 predicting social isolation at timepoint 2.

**Conclusion:** The lack of evidence of an association despite strong theoretical evidence, may be due to selection bias within the overall cohort study and the sensitivity of the outcome measures used for social isolation and cognition.

## Introduction

The consequences of unmanaged age-related hearing loss are far reaching (1), and in recent years the connection between hearing loss and cognition has been increasingly investigated (2, 3, 4). Hearing loss in midlife has been reported as the biggest modifiable risk factor (alongside high LDL cholesterol) to dementia in later life (5), thus, finding ways to prevent or delay the onset of dementia is a high priority for healthy ageing initiatives. However, the pathway connecting hearing loss and cognitive decline is yet to be established. One hypothesis to explain the relationship is a shared neuropathologic origin such as neurogenerative deterioration and inflammation (6). Another is known as the “cascade” hypothesis, which suggests that hearing loss leads to other mediating factors that exacerbate or accelerate cognitive decline (7). Social isolation may be one such mediator and has been shown to be associated with both hearing loss (8, 9) and cognitive decline (10) independently.

There has been limited research examining the role of social isolation as a mediating factor between hearing loss and cognitive function. A systematic review to investigate whether social isolation mediated hearing loss and cognition in longitudinal cohort studies (13) found that only one study out of fifteen investigated social isolation as a potential mediator, with no evidence to support its role (14). One reason for the lack of studies identified may be how the authors of the included studies within the systematic review defined social isolation and/or the demographics of the participants included in the study. Two studies that were not included in the systematic review due to their reliance on self-report over objective measures of hearing loss arrived at different conclusions. Self-reported hearing loss may be an unreliable indicator for causal pathways, with objective measures preferred to ensure better accuracy and consistency of test measures. Maharani et al (2019) investigated whether loneliness and social isolation have a role in the pathway between hearing impairment and cognitive function in older persons (11). The longitudinal study sample comprised of 8,199 people aged 50 and over. Self-reported hearing impairment was found to be negatively associated with episodic memory, and loneliness and social isolation mediated that effect within a 10-year period. The study concluded that treatments to strengthen older persons’ social networks are likely to be useful in preventing cognitive deterioration. Since only one type of cognition (episodic memory) was assessed, further studies are required to verify the role of social isolation. Another longitudinal analysis using the Chinese Longitudinal Healthy Longevity Survey investigated social isolation as a mediator between hearing loss and the Mini Mental State Examination (MMSE) as well as physical functioning outcomes (12). They found that social isolation mediated the association of self-reported hearing difficulty with cognitive functioning, but not with physical functioning. However there were limitations to the measures of social isolation: . Chen and Zhou (12) used the following four items: “When you are sick, who usually takes care of you?”; “Whom do you usually talk frequently in daily life?”; “Whom do you talk first when you need to share some of your thoughts?”; “Whom do you ask for help when you have problem or difficulties?”. If a participant answered “nobody” to any of the four questions, they were considered socially isolated, otherwise, they were deemed to have social support.

Social isolation as a phenomenon is complex and multi-faceted (15). Therefore, defining and measuring social isolation within the context of epidemiological studies is challenging (16). Some authors have used marital status and frequency of contact with relatives and friends as proxy measures for social isolation (14). Others have used a combination of measures related to social support and size of social network (17). Outcome measures such as the Medical Outcomes Survey Social Support Survey encompass a range of components related to the phenomenon of social isolation (18). This includes emotional/informational support, tangible support, affectionate support, and positive social interaction, so this type of tool to measure how isolated a person may be is more appropriate than other simplified tools.

There is a need for further longitudinal research to examine the role of social isolation in the relationship between hearing loss and cognition, using suitable measures for each of the factors ofinterest. The Hertfordshire Ageing Study (HAS) (19) was identified as a suitable dataset to address certain shortcomings of prior research. HAS employed appropriate measures for hearing, cognition, and social isolation, and utilised a representative cohort that was longitudinal in design as hearing was assessed prior to cognition and social isolation.

The following research questions were investigated in the Hertfordshire Aging Study:

Does hearing threshold predict cognitive score 10 years later?

Does hearing threshold predict social isolation score 10 years later?

## Methods

### Hertfordshire Aging Study

The HAS is a birth cohort study whose principal objective was to examine life course influences on healthy ageing (19). There were 6,803 live singletons born in North Hertfordshire between 1920 and 1930. With the help of the National Health Service Central Register, 1,428 who still lived there in 1995 were traced and 824 (58%) of the traced people agreed to a home interview. After interview, 717 men and women attended a clinic for detailed characterisation of ageing in a range of systems including hearing. Inter-and intra-observer reliability studies were carried out at regular intervals during the fieldwork to ensure comparability of measurements within and between observers.

In the HAS, participants had data collected at two points in time - roughly in 1994-5 and 2003-5. The average age of the study participants was 67 years at time point 1 and 76 years at time point 2. The first HAS follow-up (time point 1) was conducted in 1994–95 when the participants ranged in age from 63 to 73 years (mean 67). This consisted of 717 participants who underwent pure tone audiometry (0.5-4kHz) at both time points. There were 231 complete cases for hearing and cognition, and 231 complete cases for hearing and social isolation, which makes up the analysis sample.

**Figure 1.**
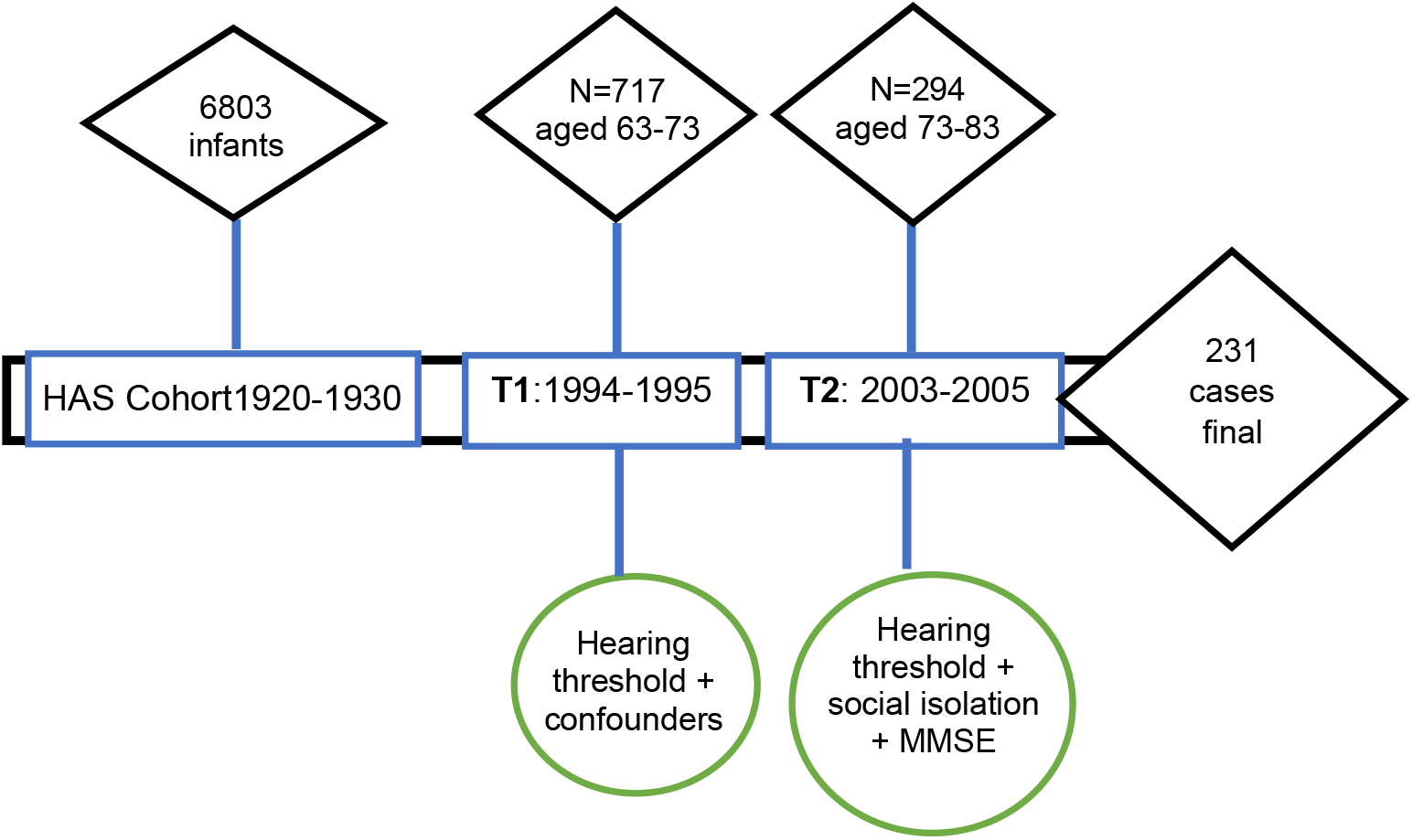
Hertfordshire Ageing Study timepoints showing participant numbers, and relevant variables measured. T1 = timepoint 1; T2 = timepoint 2; MMSE = mini mental state examination

### Hearing threshold

Hearing thresholds were assessed using pure tone audiometry. Trained researchers assessed hearing in 717 individuals at timepoint 1 and 294 individuals at timepoint 2. Audiometric thresholds were measured by air conduction at four frequencies (500, 1,000, 2,000 and 4,000 Hz). The average hearing threshold was the mean threshold value at 500, 1,000, 2,000 and 4,000 Hz by air conduction for the worse hearing ear, with higher values indicating more hearing loss. The British Society of Audiology Recommended Procedures defines normal hearing as having a hearing threshold of 20 dB or below (22). These clinical recommendations, followed by clinicians throughout the UK, are a valid way to distinguish between “normal” and “abnormal” hearing thresholds.

### Cognitive Outcomes

Cognition was assessed using the Mini-Mental State Examination (MMSE) (21) at time-point 2. This 30-point cognitive screening tool assesses the following cognitive functions: orientation, registration, attention and calculation, recall, language, and copying. The MMSE allows for a maximum score of 30. A score of less than 25 is typically seen as abnormal and indicative of possible cognitive impairment.

The MMSE, widely used in clinical practice as a dementia screening tool due to its ease of administration and comprehensive assessment of cognitive domains, is the most popular choice for assessing cognitive status (13), despite its potential disadvantage for individuals with unmanaged hearing loss due to the verbal nature of some items, requiring adequate auditory function.

### Social isolation outcome

Social isolation was assessed using the MOS Social Support Survey (18). The survey is made up of four functional support scales (emotional/informational, tangible, affectionate, and positive social interaction) and the construction of an overall functional social support index. Eight self-reported questions related to social isolations were asked, with answers given on a Likert scale from 1 (complete isolation) to 5 (no isolation). Questions included: someone you can count on to listen to when you need to talk; someone to confide in or talk to about yourself or your problems; someone to share your most private worries and fears; someone who understands your problems; someone who shows you love and affection; someone to do something enjoyable with; how often do you see children; how often do you see neighbours? Each self-reported answer was rated from 1-5 on a Likert scale with a lower score indicating greater social isolation. Social isolation was measured at time point 2 only.

### Confounding variables

Confounding variables, chosen based on the available data and the existing literature, were self-reported at timepoint 1, and were assumed to be unchanged at timepoint 2. These include: age, gender, social class, smoking status, alcohol consumption, marital status, education status, and clinical diagnoses (angina, stroke, heart attack, high blood pressure, type 2 diabetes, or depression). Self-reported social class was categorised into either professional, managerial, technical and nonmanual, or manual, partly skilled, and unskilled. This referred to a person’s own social class or their husband’s if ever married.

### Statistical analysis

The characteristics of the participants is appropriately summarised using counts and percentages for categorical variables and continuous variables summarised using either mean and standard deviation or median and inter-quartile range, depending on the data distribution. We additionally compared the characteristics of those included in the study with the non-responders using Chi-squared and Kruskal-Wallis.

For each outcome, to understand its relationship with hearing threshold, we fitted three linear regression models: an unadjusted model; a partially adjusted model (with age and sex); and a fully adjusted model (age, sex, social class, smoking status, number of alcoholic units drunk per week, marital status, years of education, diagnosed angina, stroke, heart attack, high blood pressure, type 2 diabetes, or depression).

The hearing threshold level at timepoint 1 was negatively skewed, so we have summarised it using the median and interquartile range. For the analysis, a log transformation was used to ensure normality for the linear regression analysis, before being transformed to the original scale. A complete case analysis was used ensuring that only comprehensive datasets were included in the analysis (23).

## Results

At time point two, which was conducted in 2003-2005, there was high attrition, resulting in 294 participants who had completed measures at both time points. The average age of the study participants was 67 years at timepoint 1 and 76 years at timepoint 2. The majority (58.8%) were male, and were current smokers (10.2%) or ex-smokers (53.1%) (Table 1). Most of the participants were married (74.1%), drank 10 units or less of alcohol per week (49%) or no alcohol (34%). Of the clinical diagnoses, high blood pressure was the most prevalent (30.2%), with depression (13.3%) and heart attack (9.6%) the next most prevalent.

**Table 1.**
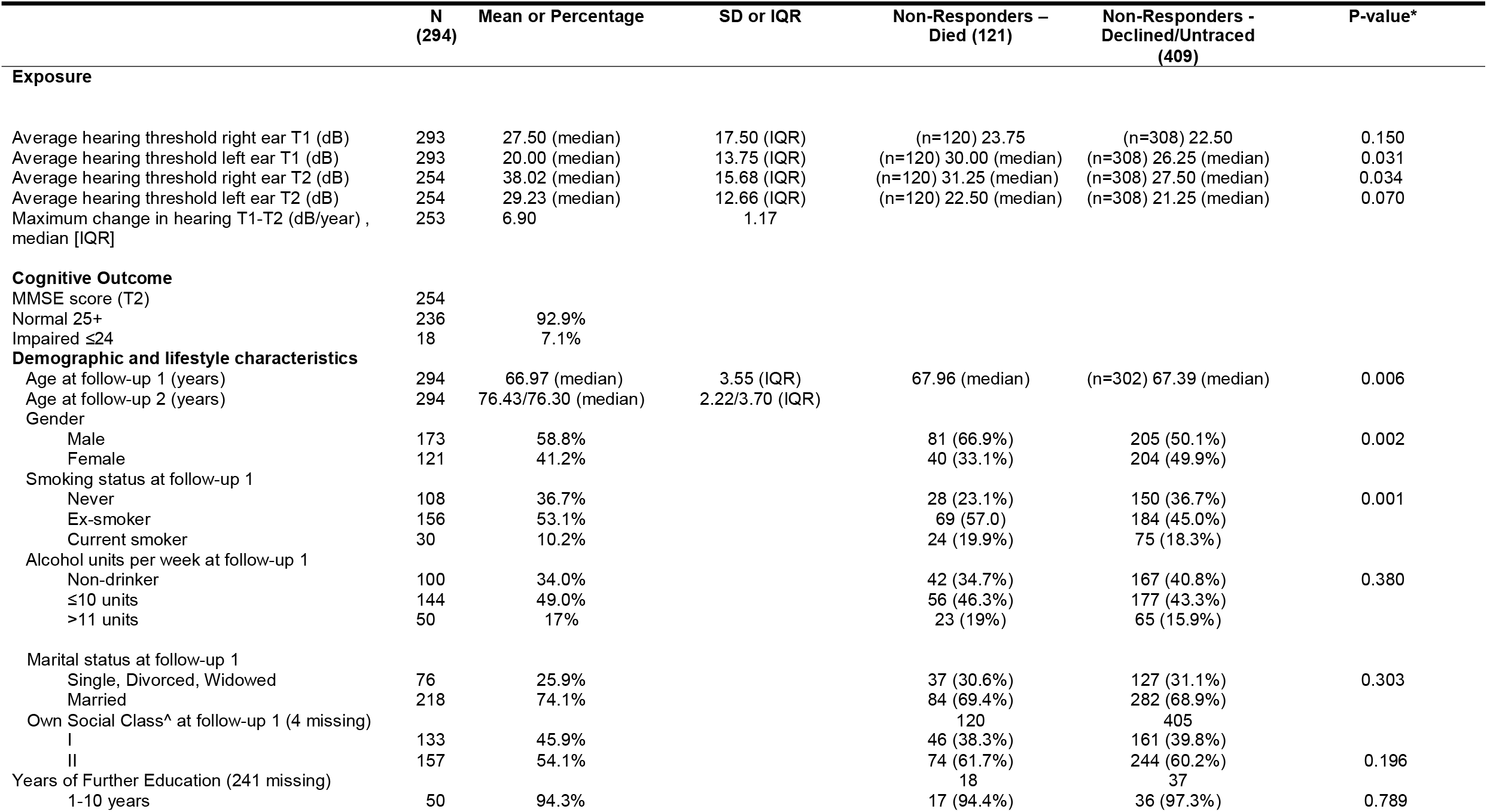
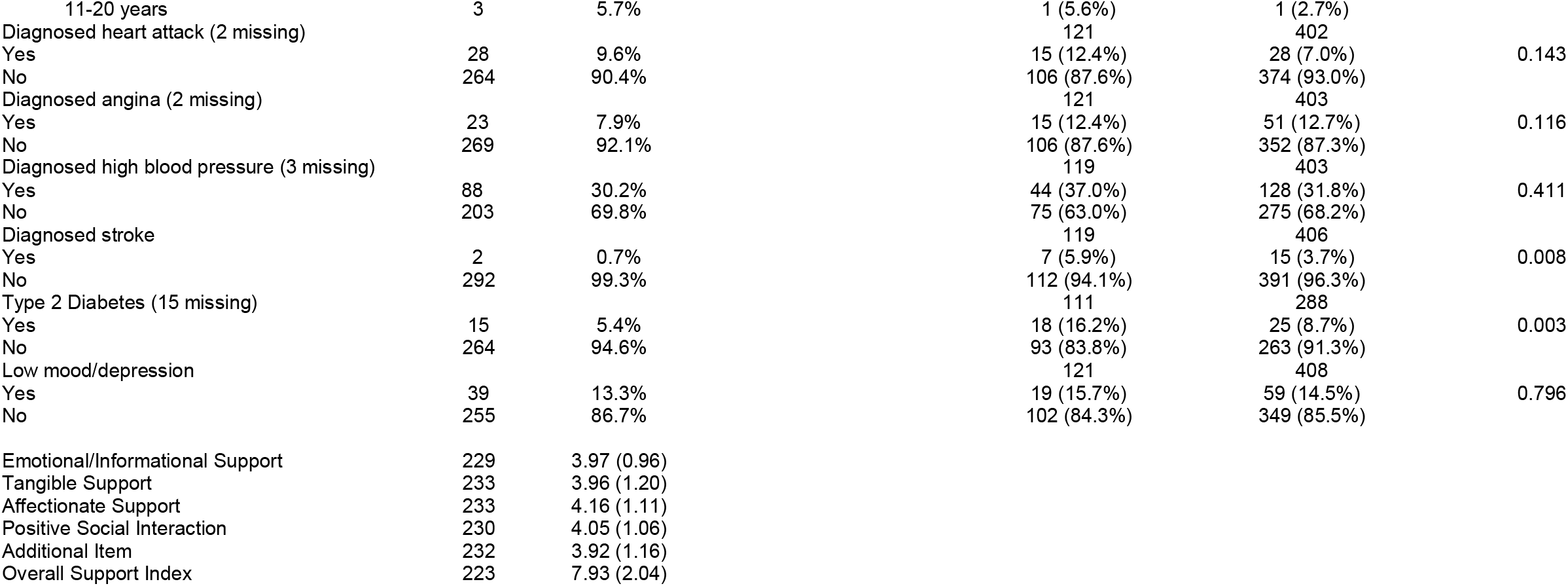
Population characteristics of included sample compared with non-responders.

When comparing non-responders (those who died before T2 and those who declined/untraced) to the study participants, the non-responders were generally: older age, male gender, worse average hearing threshold, current smoking status at time point 1, drinking >11 alcohol units per week, a marital status of single/divorced/widowed, lower social class, diagnosis of stroke and type 2 diabetes.

The median worse-ear hearing threshold level (referred as “hearing threshold”) was 27.50 dB HL at time point 1, worsening to 38.02 dB HL at time point 2. Therefore, there was an average decline of 10.5 dB. The rate of change between time points 1 and 2 in hearing threshold in dB per year was a maximum of 6.9dB, a mean (SD) of 0.76 (1.17).

### Hearing loss and cognition

Of the 294 in the final included sample, 254 participants had completed the MMSE data, and 231 had completed the social isolation survey. For MMSE score at time point 2, 92.9% of participants scored 25 or more (normal range).

While the unadjusted analysis in table 2 (model 1) showed a relationship between hearing threshold and later cognitive score, age- and sex-adjusted (model 2) and fully adjusted models (model 3) suggested no evidence of a significant association between hearing threshold and later cognitive score. The effect size for hearing threshold and later cognitive score in the unadjusted model was - 1.476 95% CI (-2.992, 0.039), *p* =0.056. For the model adjusting for age and sex, the effect size was - 1.067 95% CI (-2.586, 0.453), *p*= 0.168. The effect size for the fully adjusted model was -0.923 95% CI (-2.471, 0.625), *p* = 0.241. This tells us that an increase in hearing threshold results in a decline in cognition. The results are not statistically significant and may be explained, at least in part, by age, sex, and other confounders such as social class, smoking status, alcohol consumption, marital status, education status, and clinical diagnoses (angina, stroke, heart attack, high blood pressure, type 2 diabetes, or depression). The decrease in effect size with increased adjustment suggests the influence of the aforementioned confounding variables on observed associations.

**Table 2.** Regression coefficients for hearing threshold (exposure) at time point 1 and cognitive scores (outcome) at time point 2.

| **Linear Regression Models** | **Change in MMSE per dB HL increase** | **95% CI (Lower, Upper)** | **P-value** |
| --- | --- | --- | --- |
| Model 1 – Univariable | -1.476 | -2.992, 0.039 | 0.056 |
| Model 2 – Adjusted for age and sex | -1.067 | -2.586, 0.453 | 0.168 |
| Model 3 – Fully adjusted | -0.923 | -2.471, 0.625 | 0.241 |

**Table 3.**
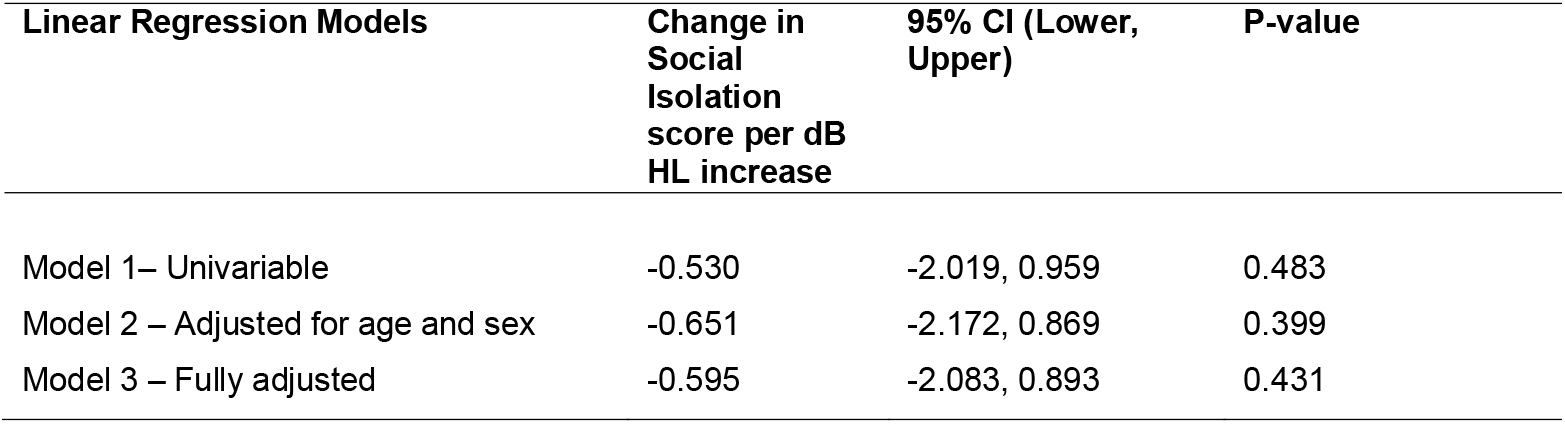
Regression coefficients for hearing threshold at time point 1 and social isolation scores at time point 2.

### Hearing loss and social isolation

The effect size for hearing threshold and later social isolation in the unadjusted model was -0.530, 95% CI (-2.019, 0.959), p = 0.483 (table 6). For the model adjusting for age and sex, the effect size was -0.651, 95% CI (-2.172, 0.869), p = 0.399. The effect size for the fully adjusted model was -0.595, 95% CI (-2.083, 0.893), p = 0.431. These negative effect sizes suggest a potential inverse relationship between hearing loss and social isolation—meaning that as hearing loss increases, there may be a trend toward lower levels of social isolation. However, because the confidence intervals for all models are wide and include zero, and the p-values are well above the conventional threshold for significance (p < 0.05), there is insufficient statistical evidence to support a meaningful association between hearing loss and social isolation. In other words, the observed estimates might be due to random variation rather than a true relationship.

## Discussion

This study investigated the longitudinal relationship between hearing threshold and later cognitive function, as well as hearing threshold and later social isolation, in community-dwelling older adults. Over a 10-year period, the cohort’s average hearing threshold worsened by 10.5 dB. However, unlike prior research findings (24, 25, 26), our analysis revealed no statistically significant associations between hearing loss and cognitive score or between hearing loss and social isolation score. Although no significant associations were found, the direction of these exposure-outcome relationships was inverse, in line with other studies (27, 28).

Several factors may explain our findings which are different to those of previous studies. Our analysis controlled for thirteen confounders, including age. A recent cohort study also found no significant effect of hearing loss on cognitive decline after adjusting for age (29), underscoring the importance of accounting for confounders and employing outcome measures sensitive to subtle, long-term changes. Additionally, evidence suggests that statistically significant findings may be over-represented in the literature (30,31) with a survey of 4,600 publications indicating a trend of publication bias, with journal editors favouring significant results to enhance impact factors (31). This bias makes challenging prevailing research trends difficult, contributing to a lack of published data on non-associations, such as between hearing loss and cognition.

Our study’s relatively small sample size, compared to previous studies (3, 24, 25), may have led to underpowering, thus limiting our ability to detect associations. Although the dataset was fixed after data collection, attrition between the study’s time points contributed to a smaller final sample, with missing predictor data further reducing the effective sample size. Participants who died or declined to continue likely differed in key characteristics, including accelerated cognitive decline or increased social isolation, impacting our findings.

Moreover, our outcome measures may lack sensitivity for this sample. Epidemiological data might not capture nuanced patterns in hearing-cognition or hearing-social isolation relationships as effectively as individual-level observations. For instance, the MMSE may not sensitively track cognitive changes in older adults (32), and those absent at time point 2 may have experienced more severe decline or isolation, skewing the sample.

The hearing threshold decline (10.5 dB) over 10 years is not considered clinically significant, as a decline of 15 dB or more at certain frequencies is generally needed to denote clinical relevance in older adults (33, 34). Furthermore, the community-dwelling participants had considerable control over their acoustic environment, potentially compensating for mild hearing changes. Most participants also maintained MMSE scores within the normal range, suggesting minimal cognitive impact from the observed hearing decline.

While the sample characteristics resemble nationally representative data for older adults in England and Wales, they may not fully represent older adults with concurrent hearing loss, cognitive decline, and social isolation. The sample included community-dwelling individuals in good health, limiting the generalisability of findings to more vulnerable populations. If levels of cognitive impairment and social isolation are broadly low in the analysis sample, then there is just not the variation in these outcomes to observe differences in these outcomes according to hearing threshold. Future studies should consider inclusive criteria to avoid selection and attrition biases that could distort exposure-outcome relationships (35).

This study has several strengths. Hearing was measured objectively using pure-tone audiometry, reducing inaccuracies associated with self-reported hearing data, which may have led to biased results in hearing-related research. The MOS social survey, a validated instrument, was employed to assess social isolation comprehensively, avoiding reliance on proxy measures such as marital status or living arrangements, which may be inadequate indicators of isolation.

Our study does, however, have limitations related to potential selection bias. The high attrition rate, with non-respondents differing significantly in age, smoking status, hearing threshold, and cognitive scores, could mean that those with poorer outcomes were excluded from the analysis. Selection bias may therefore lead to misleading estimates of association.

Additionally, the hearing changes observed over the study’s intervals were modest, and more frequent follow-ups over longer timeframes may improve the power to detect associations. Our study design did not permit mediation analysis of social isolation on cognitive decline, but this warrants exploration in future studies to better understand the interplay between hearing, cognition, and social relationships.

In conclusion, while this study contributes to the literature on hearing loss, cognitive decline, and social isolation, it highlights the need for more sensitive cognitive and social isolation measures. Longitudinal research using well-defined inclusion criteria and sensitive outcome measures, such as cross-lagged panel or latent difference score models, may advance understanding in this area. Given the ageing population, healthy ageing initiatives—including regular hearing and cognitive screening and socially interactive programmes—will be essential to mitigating cognitive decline and social isolation among older adults.

## Data Availability

The data can be accessed via the MRC Lifecourse Epidemiology centre at University of Southampton.

## Declarations

## Ethics approval and consent to participate

Ethics approval not required due to secondary data analysis. Original consent to participate and ethics approval was sought by the Hertfordshire Ageing Study research group.

## Consent for publication

The data are not published elsewhere. There is full consent for publication according to the data sharing agreement with Hertfordshire Ageing Study research group.

## Availability of data and materials

The data can be accessed via the MRC Lifecourse Epidemiology centre at University of Southampton.

## Authors’ contributions

N.D. requested the data, completed analysis, wrote the main manuscript text and prepared all tables.

A.H. edited the manuscript text and supported data analysis and interpretation.

J.M. checked the accuracy of the statistical analysis.

H.P. edited the manuscript text.

All authors reviewed the manuscript.

## Acknowledgements

Hertfordshire Ageing Study participants and researchers.

Dr Leo Westbury (statistician at MRC Lifecourse Epidemiology Centre) Professor Avan Aihie Sayer (Hertfordshire Ageing Study Principal Investigator)

